# Circulating cell-free messenger RNA secretome characterization of primary sclerosing cholangitis

**DOI:** 10.1101/2022.08.22.22278964

**Authors:** Naga Chalasani, Raj Vuppalanchi, Craig Lammert, Samer Gawrieh, Jerome V. Braun, Jiali Zhuang, Arkaitz Ibarra, David A. Ross, Michael Nerenberg, Stephen R. Quake, John J. Sninsky, Shusuke Toden

## Abstract

**BACKGROUND & AIMS:** Primary sclerosing cholangitis (PSC) is a rare chronic cholestatic liver disease characterized by multi-focal bile duct strictures. To date, underlying molecular mechanisms of PSC remain unclear and therapeutic options for PSC patients are limited. We performed cell-free messenger RNA (cf-mRNA) next generation sequencing (RNA-Seq) to characterize the circulating transcriptome of PSC and non-invasively investigate potentially bioactive signals that are associated with PSC.

**METHODS:** Serum cf-mRNA profiles were compared among 50 individuals with PSC, 20 healthy controls, and 235 individuals with non-alcoholic fatty liver disease (NAFLD). Tissue and cell type-of-origin genes that are dysregulated in subjects with PSC were evaluated. Subsequently, diagnostic classifiers were built using PSC dysregulated cf-mRNA genes.

**RESULTS:** Differential expression analysis of the cf-mRNA transcriptomes of PSC and healthy controls resulted in identification of 1407 dysregulated genes. Furthermore, differentially expressed genes between PSC and the liver diseases (NAFL and Non-Alcohol Steatohepatitis (NASH)) or healthy controls shared common genes known to be involved in liver pathophysiology. In particular, genes from liver- and specific cell type-origin, including hepatocyte, hepatic stellate cells and Kupffer cells, were highly abundant in cf-mRNA of subjects with PSC. Gene cluster analysis revealed that liver-specific genes dysregulated in PSC form a distinct cluster which corresponded to a subset of the PSC subject population. Finally, we developed a cf-mRNA classifier using liver-specific genes which discriminated PSC from healthy control subjects using gene transcripts of liver origin.

**CONCLUSIONS:** Blood-based whole transcriptome cf-mRNA profiling revealed high abundance of liver-specific genes in PSC subject sera which may be used to diagnose PSC patients. We identified several unique cf-mRNA profiles of subjects with PSC. These findings may also have utility for non-invasive molecular stratification of subjects with PSC for pharmacotherapy safety and response studies.

**Graphical Abstract:** 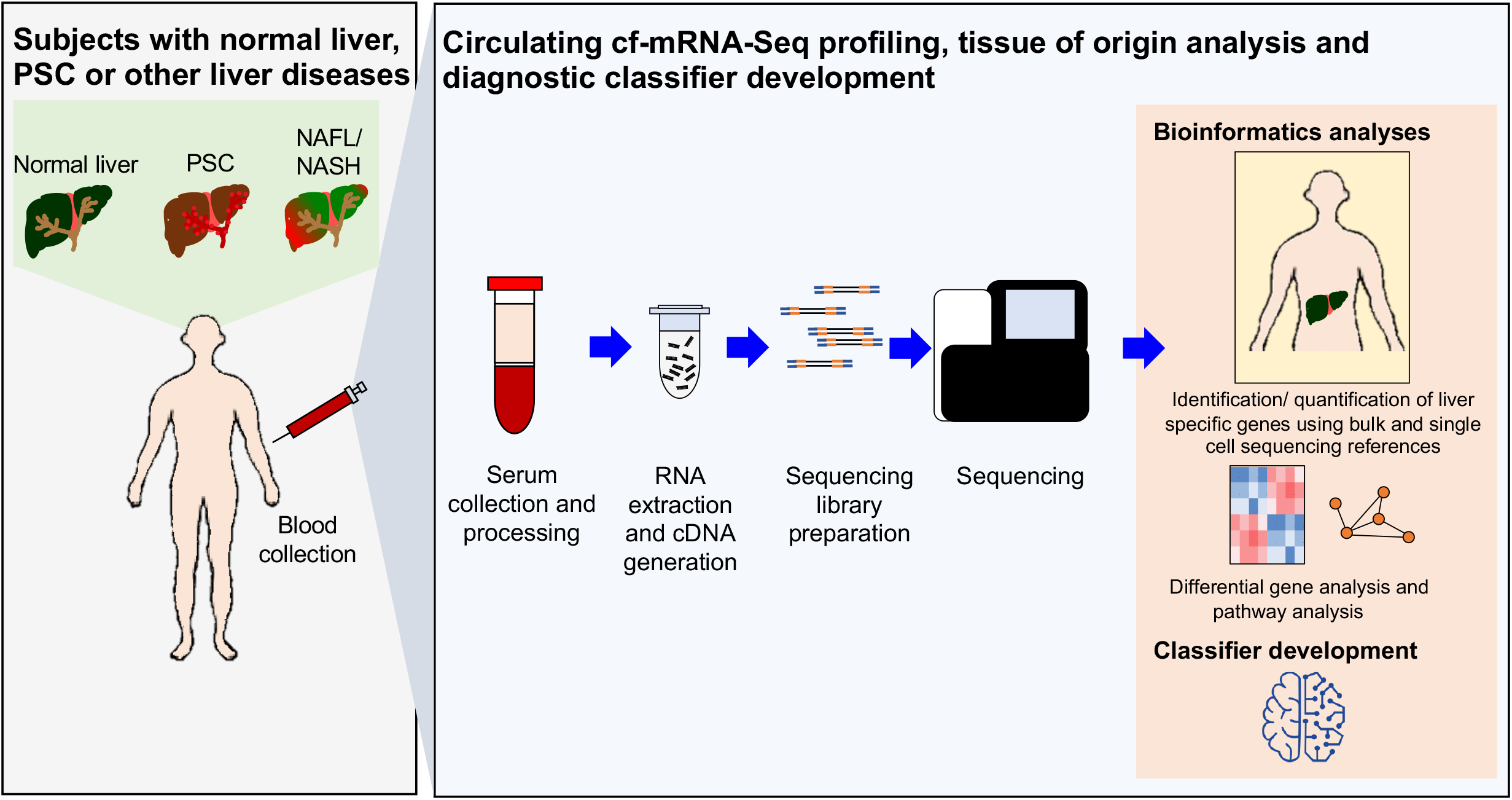

## INTRODUCTION

Primary sclerosing cholangitis (PSC) is a rare chronic cholestatic syndrome characterized by inflammation and fibrosis of the intra- and extra-hepatic bile duct leading to multifocal biliary strictures. PSC is known to be associated with inflammatory bowel disease (IBD) with two-thirds of the PSC patients in the United States concurrently diagnosed with IBD *(1, 2)*. Furthermore, PSC is a premalignant condition associated with increased risk for cholangiocarcinoma and colorectal cancer *(3)*. With limited therapeutic options and a lack of proven monitoring strategies, there are significant clinical unmet needs for patients with PSC *(1)*.

Currently, magnetic resonance imaging (MRI) is the non-invasive imaging modality of choice for diagnosing suspected PSC *(4, 5)*. Although liver histological findings correlate with a PSC diagnosis, histological changes are generally indicative of some form of biliary disease, but not specific to PSC *(6)*. Therefore, liver biopsy for PSC diagnosis is typically performed on a subset of patients suspected of having the small-duct variant of PSC, in whom MRI does not have sufficient imaging resolution to characterize the microscopic and fine branch bile ducts *(7)*. Considering that the majority of subjects with small-duct PSC as well as a significant portion of subjects with large-duct PSC are asymptomatic *(8)*, there is a need for a highly accessible, simple blood-based non-invasive diagnostic test for PSC to circumvent delays in disease diagnosis. Moreover, one of the major hindrances for PSC therapeutic development is lack of clear understanding of PSC etiology, progression and pathogenesis. While recent characterization of immune cells in the bile duct of subjects with PSC *(9)* and evaluation of proteomic and microRNA characteristics of exosomes derived from subjects with PSC *(10)* have revealed previously unknown molecular characteristics of PSC, unlike other liver diseases, molecular characterization of PSC remains limited. Hence, there is a critical need to improve the understanding of underlying molecular mechanisms of PSC in order to identify potential therapeutic targets for new therapeutic options and surveillance management strategies to inform disease progression.

Liquid biopsy has emerged as a viable non-invasive alternative to surgical biopsies for diagnosis of various diseases *(11-13)*. Recent advancements in next generation sequencing (NGS) have accelerated the incorporation of circulating nucleic acids in the development of non-invasive disease biomarkers *(14, 15)*. In particular, we have developed methods for quantification of circulating cell-free messenger RNA (cf-mRNA) using NGS platforms for RNA-Seq *(16-19)*. Through utilization of well-characterized human tissue RNA sequencing databases, the tissue and cell type of origin of circulating mRNA transcripts can be determined *(19-21)*. Recently, we conducted cf-mRNA transcriptome profiling of subjects with non-alcoholic fatty liver disease (NAFLD) using cf-mRNA RNA-Seq. We identified genes that are dysregulated in subjects with NAFLD compared to healthy controls and developed diagnostic classifiers for subjects with advanced liver fibrosis *(18)*. Here, we performed cf-mRNA RNA-Seq profiling on serum specimens obtained from 50 PSC and 20 healthy control subjects. We compared the cf-mRNA profiles of PSC patients and that of healthy controls and identified transcriptional profiles distinct to PSC. Furthermore, the gene-expression profile of subjects with PSC also substantially differed from those with NAFLD and NASH in our prior study *(18)*. We discovered that subjects with PSC had remarkably elevated levels of liver tissue- and liver cell type-specific genes in the circulation, independent of their IBD status. Cluster analysis revealed that a subset of subjects with PSC had a high abundance of liver-specific gene transcripts. Finally, we developed diagnostic cf-mRNA classifiers for PSC, which accurately discriminated subjects with PSC from healthy controls. Comparison of subjects with PSC and our prior NAFLD and NASH study permitted an investigation of the common cf-mRNA signals. Our study highlights the potential utility of cf-mRNA-based profiling for non-invasive molecular characterization of subjects with PSC, identification of potential therapeutic targets and development of non-invasive biomarkers for PSC.

## RESULTS

### Technical Performance of cf-mRNA RNA-Seq Assay in Human Serum Specimens

We demonstrated previously that our cf-mRNA RNA-Seq machine learning platform is highly sensitive and reproducible for Alzheimer’s disease, transplant oncology and NAFLD *(16-18)*. Considering that the sequencing data will influence subsequent downstream bioinformatic analyses, generation of reliable RNA-Seq data is critical (Figure 1A). We evaluated the key metrics for library fidelity to ensure the quality of the libraries that we generated in the study. We assessed the number of protein coding genes identified per sample to assess the sensitivity of our assay. We identified 9,234 protein coding genes per sample (mean value, TPM > 5) (Supplementary Fig. 1A). Furthermore, we showed high correlation between the observed versus expected number of extracellular RNA consensus consortium (ERCC) reference material for the libraries generated for the study (mean Pearson correlation coefficient = 0.95) (Supplementary Fig. 1B and 1C), confirming the quality of our library preparation method. Collectively, these data are consistent with our previous studies *(16-18)* and demonstrate the reproducibility of the assay.

**Figure 1:**
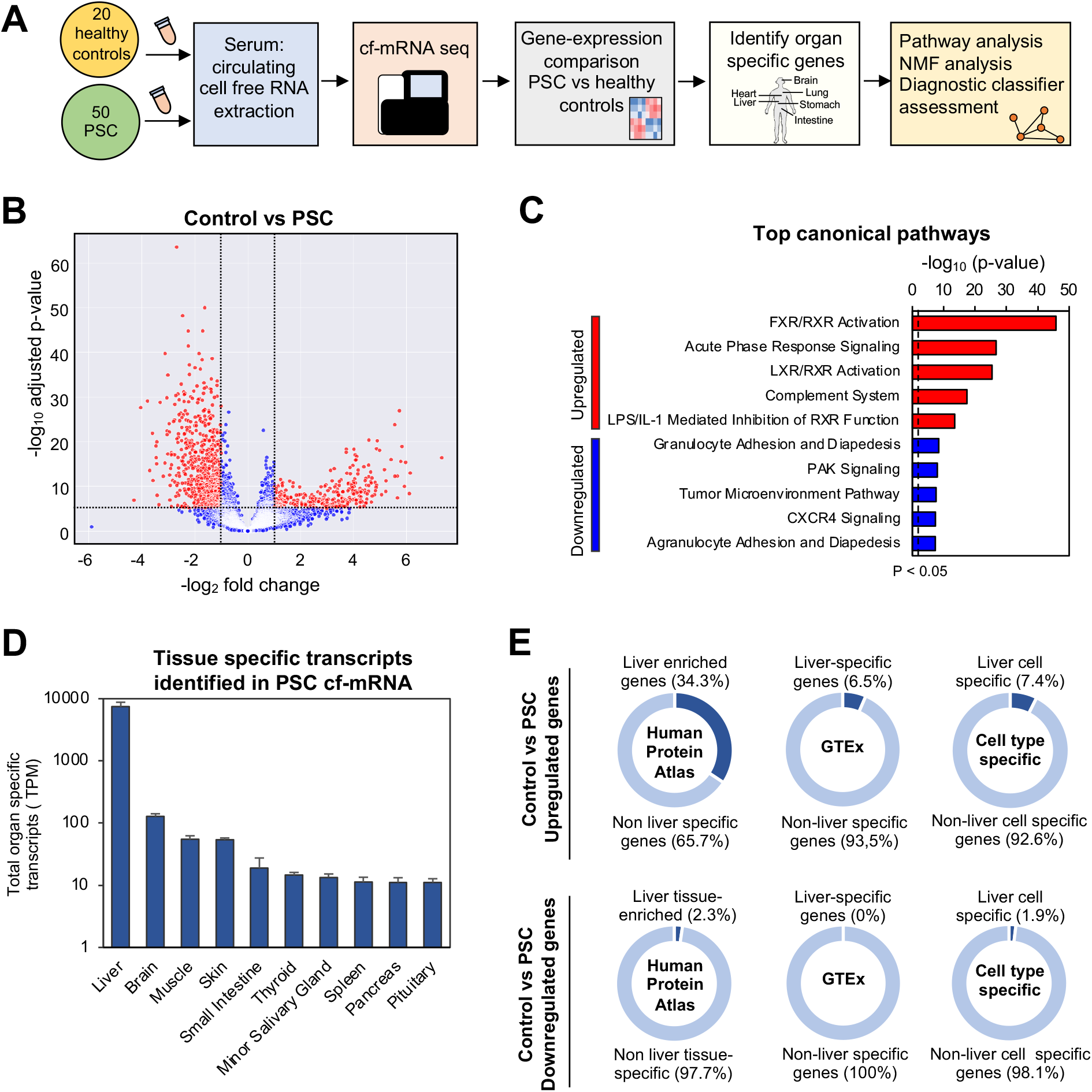
Identification of high levels of liver-specific genes in cf-mRNA of subjects with PSC. (A) Schematics of the study design. (B) A volcano plot showing the dysregulation of cf-mRNAs in subjects with PSC compared to that of healthy controls. (C) Top IPA canonical pathways for genes that are dysregulated in PSC, upregulated and downregulated genes are used as input. (D) Quantification of tissue specific transcripts identified in cf-mRNA of subjects with PSC (Mean ± SEM). (E) Quantification of genes that are associated with liver in genes that are differentially expressed in subjects with PSC compared to healthy controls (upregulated genes, top; downregulated genes, bottom). Reference data sets used to calculate the liver tissue and cell type specificities are: Human Protein Atlas (HPA) (left), GTEx (middle) and PanglaoDB (right).

### Identification of elevated levels of liver-specific transcripts in subjects with PSC

In order to examine the cf-mRNA transcriptomic profile of subjects with PSC compared to that of healthy individuals, we evaluated cf-mRNA sequencing data generated from sera of 50 PSC with PSC compared to 20 healthy controls using differential expression analysis to identify circulating transcripts that are dysregulated in the subjects with PSC. Using the following cut-off criteria: adjusted p < 0.05 and fold change > 2, we identified 1,742 dysregulated genes in the circulation of subjects with PSC, 912 upregulated and 830 downregulated (Figure 1B). (The terms “upregulated” and “downregulated” are used to describe changes in the number of RNA molecules in the circulation of PSC compared to control subjects). Next, using the differentially expressed genes between PSC and healthy controls, we performed Ingenuity Pathway Analysis (IPA) gene-enrichment analysis (Qiagen) to examine pathways that are dysregulated in the cf-mRNA transcriptome of subjects with PSC. The pathway analysis identified canonical pathways that are involved in liver metabolism, such as FXR/RXR activation and acute phase response signaling, as the most significantly enriched pathways for upregulated genes, while granulocyte adhesion and PAK signaling were the most significant pathways identified in the downregulated genes (Figure 1C). Furthermore, additional pathway analyses using alternative pathway analysis (Gene Ontology, KEGG and Reactome) indicated that the genes that are dysregulated in cf-mRNA of PSC subjects are involved in acid metabolism (Gene Ontology) and the complement cascade (KEGG and Reactome) (Supplementary Fig. 2A, 2B and 2C), pathways that are known to be associated with PSC *(22, 23)*. Considering that the pathways that we identified using upregulated genes in IPA had substantially higher adjusted p-values compared to those of downregulated genes, the upregulated pathways may be more likely to have higher effect sizes (Figure 1C). Since we have identified several liver metabolism associated pathways in PSC, we evaluated the distribution of tissue-specific genes in the whole transcriptome of subjects with PSC and healthy controls using a criterion that we developed using the Genotype-Tissue Expression (GTEx) project as a reference *(24)* (See Materials and Methods). For both PSC and healthy controls, liver-specific gene transcripts were the most abundant followed by brain-specific gene transcripts (Figure 1D and Supplementary Fig. 3A). Moreover, PSC subjects had substantially more liver-specific gene transcripts compared to healthy controls (12.4-fold, 7426 ± 1354 vs 597 ± 153 respectively, *p* < 0.001). Next, we used three independent annotations to evaluate the portion of liver genes that are differentially expressed between subjects with PSC and healthy controls (Figure 1E, Supplementary Fig. 3B). Approximately a third (34.3%) of genes that are differentially expressed in PSC subjects were liver-specific, using the Human Protein Atlas (HPA) as the reference *(25)*. Furthermore, we identified 6.5% liver-specific (GTEX as the reference *(24)*) and 7.4% liver cell-type specific gene transcripts using PanglaoDB as the reference *(26)*). We also evaluated the abundance of liver gene transcripts using those downregulated in PSC subjects for comparison (Figure 1E). Collectively, these data indicated that the proportion of differentially expressed gene transcripts in subjects with PSC varied from 6.5-34.3% depending on the reference used.

In addition, we conducted differential expression analysis between PSC and NAFL and NASH to compare the dysregulated cf-mRNA transcripts between these two liver diseases (Supplementary Fig. 4A). We used cf-mRNA-Seq data that we generated previously *(18)*. Samples used to generate NAFL and NASH cf-mRNA-Seq data were collected at the same institution (Indiana University) and processed using the same methods as the current study *(18)*. IPA pathway analysis confirmed that similar liver metabolism associated genes were dysregulated in PSC when compared to NAFL and NASH (Supplementary Fig. 4B) but substantially higher levels of liver-specific genes were upregulated in cf-mRNA of subjects with PSC compared to subjects with NAFL and NASH (Supplementary Fig. 5). Collectively, our data indicate that subjects with PSC appear to have elevated levels of transcripts that are derived from liver in the cf-mRNA secretome.

### Distribution of liver cell types in cf-mRNA of PSC subjects

In order to further evaluate the gene-expression profile of cf-mRNA of subjects with PSC, we assessed the distribution of liver cell types for genes that were upregulated in these subjects using a reference that is derived from single cell sequencing datasets (PanglaoDB) *(26)* (Figure 2A). The majority of upregulated liver genes in subjects with PSC were hepatocyte-associated (78%), while other liver cell types that we identified include: hepatoblasts (bi-potential cells which differentiate into hepatocytes or biliary epithelial cells), hepatic stellate cells, Kupffer cells (resident liver macrophages) and cholangiocytes (Figure 2A). The number of liver-specific gene transcripts (Figure 2B) as well as the total number of liver cell-specific transcripts (Figure 2C) were abundant in subjects with PSC compared to healthy controls. The comparison of the individual liver cell-type specific gene transcripts between PSC and healthy control subjects indicated that subjects with PSC had a higher abundance cf-mRNA from hepatocyte, hepatoblasts and cholangiocytes than hepatic stellate cells and Kupffer cells (Figure 2D). To further evaluate the tissue- and cell-type specificity of key dysregulated genes in PSC, we compared the top 10% of genes that are upregulated in subjects with PSC to those of healthy controls (Figure 2E, Supplementary Fig. 6A). The proportion of liver genes in the top 10% upregulated genes were substantially higher than that of total upregulated genes (Figure 2E). We next used IPA enrichment analysis to investigate the key upstream regulator genes of the upregulated genes in PSC subjects (Figure 2F). We identified HNF1A, HNF4A, PPARA and NR1H4 genes as the key upstream transcriptional regulators. While NR1H4 and PPARA are gene transcripts are differentially expressed in hepatic stellates cells, HNF1A and HNF4A have been shown to be associated with regulation of liver fibrosis *(27-29)*.

**Figure 2:**
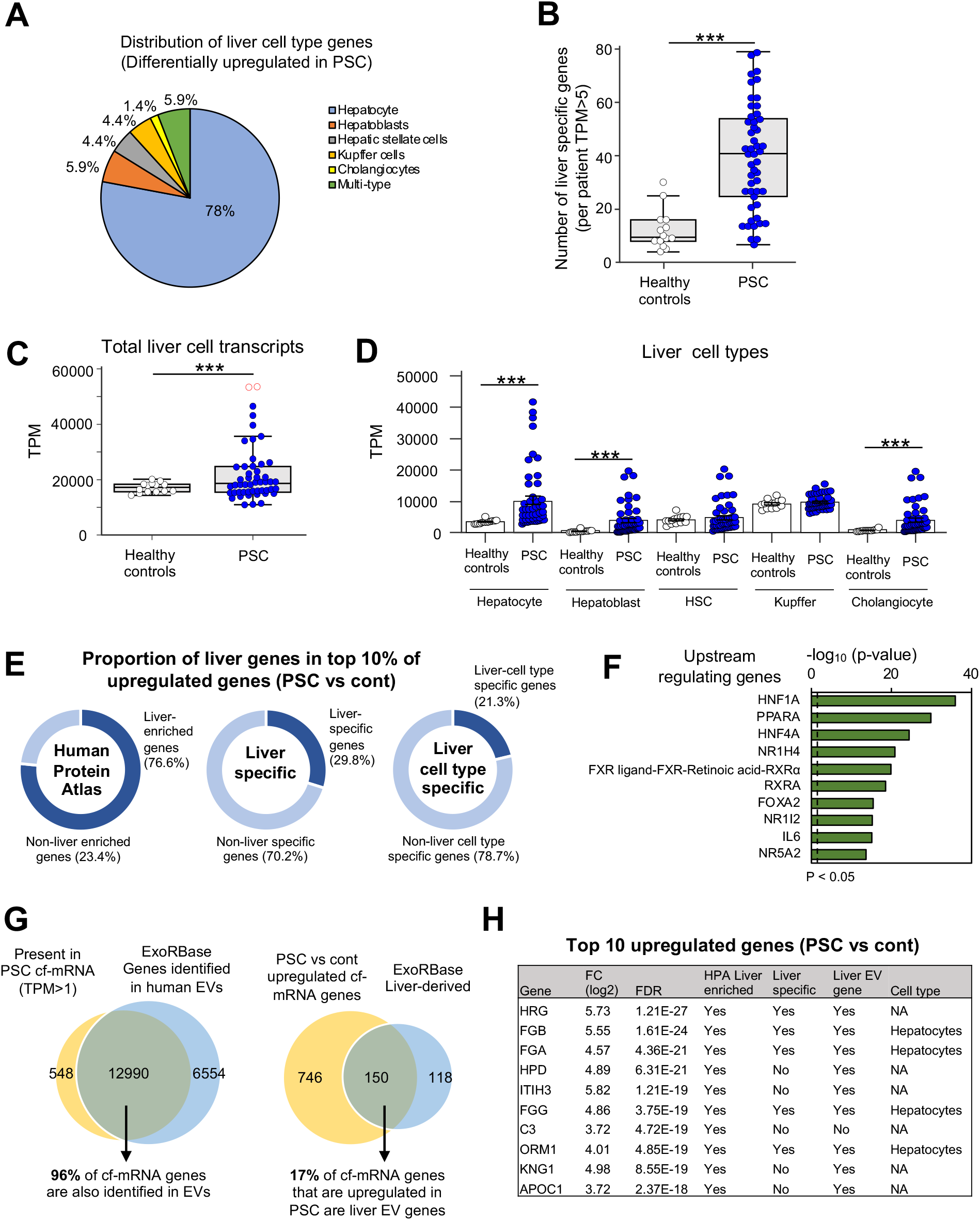
Distribution of liver specific genes. (A) Distribution of liver cell type specific genes in genes that are differentially upregulated in subjects with PSC compared to healthy controls. (B) The total number of liver tissue specific genes per sample. Subjects with PSC left, healthy controls right. (C) The total number of liver cell specific genes per samples. Subjects with PSC left, healthy controls right. (D) The abundance of liver cell specific genes per sample based on cell types. (E) Quantification of genes that are associated with liver in genes in the top 10% upregulated genes in subjects with PSC compared to healthy controls. Reference data sets used to calculate the liver tissue and cell type specificities are: Human Protein Atlas (HPA) (left), GTEx (middle) and PanglaoDB (right). (F) IPA was used to evaluate the upstream regulating genes (PSC genes that were upregulated compared to downregulated genes are used as the inputs). (G) The overlap between genes that were identified in cf-mRNA of subjects with PSC (TPM > 1) and genes identified in human extracellular vesicles (EVs) (ExoRBase used as the reference). (H) A table summarizing the liver associated molecular characteristics of the top 10 most upregulated genes in cf-mRNA of subjects with PSC compared to healthy controls.

Cf-mRNA is protected from RNase digestion due to embedding in extracellular vesicles with lipid bilayers, membraneless particles *(30, 31)* as well as other ribonucleoprotein complexes. In particular, studies have begun to survey the cfRNA in membrane bound extracellular vesicles such as exosomes, microvesicles and apoptotic bodies *(32)*. We, therefore, examined the overlap between gene transcripts that were identified in cf-mRNA of subjects with PSC (TPM > 1) with the gene transcripts that were identified in human extracellular vesicles using the ExoRBase database *(33)*. Ninety-six percent of gene transcripts that were found in cf-mRNA of PSC subjects were also present in the ExoRBase database (Figure 2G). Furthermore, 17% of the liver-specific gene transcripts upregulated in subjects with PSC subjects were listed in the ExoRBase database (Figure 2G). Finally, we investigated the molecular characteristics of the genes that are the 10 most highly upregulated in cf-mRNA in PSC subjects compared to healthy controls (Figure 2H). All 10 gene transcripts were liver related- (HPA) while 9/10 of genes were in the ExoRBase database, 5/10 genes were liver-specific (GTEx) and 4/10 genes were of specific hepatocyte origin (PanglaoDB). These data suggest that the majority of most highly upregulated cf-mRNA gene transcripts are of liver origin. Moreover, to further confirm our findings, we assessed the top 10 upregulated genes between cf-mRNA of subjects with PSC to those with NAFL and NASH (Supplementary Fig. 7). Consistent to the comparison to healthy controls, we found abundance of liver tissue and cell type specific genes in subjects with PSC when compared to those with NAFL and NASH. However, none of top 10 downregulated genes were associated with liver for PSC v healthy controls, PSC v NAFL and PSC v NASH (Supplementary Fig. 7). Collectively, our data demonstrated that the liver origin gene transcripts are abundant in the cf-mRNA secretome of subjects with subjects with PSC.

### Cf-mRNA expression levels of liver cell type specific genes in PSC subjects

To investigate whether the overall cf-mRNA gene expression profile of subjects with PSC is substantially different from NAFLD and NASH, we plotted the fold change of all genes for PSC, NASH and NAFL using genes from healthy controls as the reference (Figure 3A). The distribution of fold changes of genes compared to healthy controls increased according to the severity of the liver disease (NAFL < NASH < PSC). Furthermore, when we specifically assessed the liver-specific genes in subjects with PSC compared to those of healthy controls, all of the liver-specific gene transcripts were more abundant in this group than in healthy controls (Figure 3B). Considering that the hepatic stellate cells have been associated with a progression of liver fibrosis and the cross talk between various liver cell types in liver disease *(34, 35)* (Figure 3C), we next evaluated the individual expression levels of several key liver cf-mRNA gene transcripts. The expression levels of HRG along with hepatocyte-associated genes, FGB and FGA, were highly elevated in a subset of subjects with PSC but these genes were rarely expressed in healthy controls (Figure 3D). NR1H4, also known as FXR, is a hepatocyte and hepatic stellate cells associated gene. A double-blinded clinical study has shown that FXR agonist improved markers of cholestasis and liver injury in subjects with PSC *(36)*. PPAR is another hepatocyte and hepatic stellate cell associated gene that is targeted for multiple liver diseases to suppress fibrosis *(29)*. Both NR1H4 and PPAR were also identified as key upstream regulator of genes that are dysregulated in PSC and were elevated in a subset of subjects with PSC (Figure 3E). Kupffer cells play a critical role in both hepatic and systemic response to pathogens and have been known to be elevated in PSC *(37)*. Consistently we observed increased levels of Kupffer cells associated genes, C1QA, C1QB and C1QC in the cf-mRNA of subjects with PSC (Figure 3F). Finally, we examined the expression level of cholangiocyte associated genes, LGALS4 and HNF1A, upstream regulators which are involved in liver fibrosis *(28)*. While both genes were expressed at low levels in healthy controls, a subset of subjects with PSC had elevated levels of these cf-mRNA gene transcripts (Figure 3G). Collectively, these data show that various genes that are known to be involved in pathophysiology of PSC are highly upregulated in the cf-mRNA of subjects with PSC.

**Figure 3:**
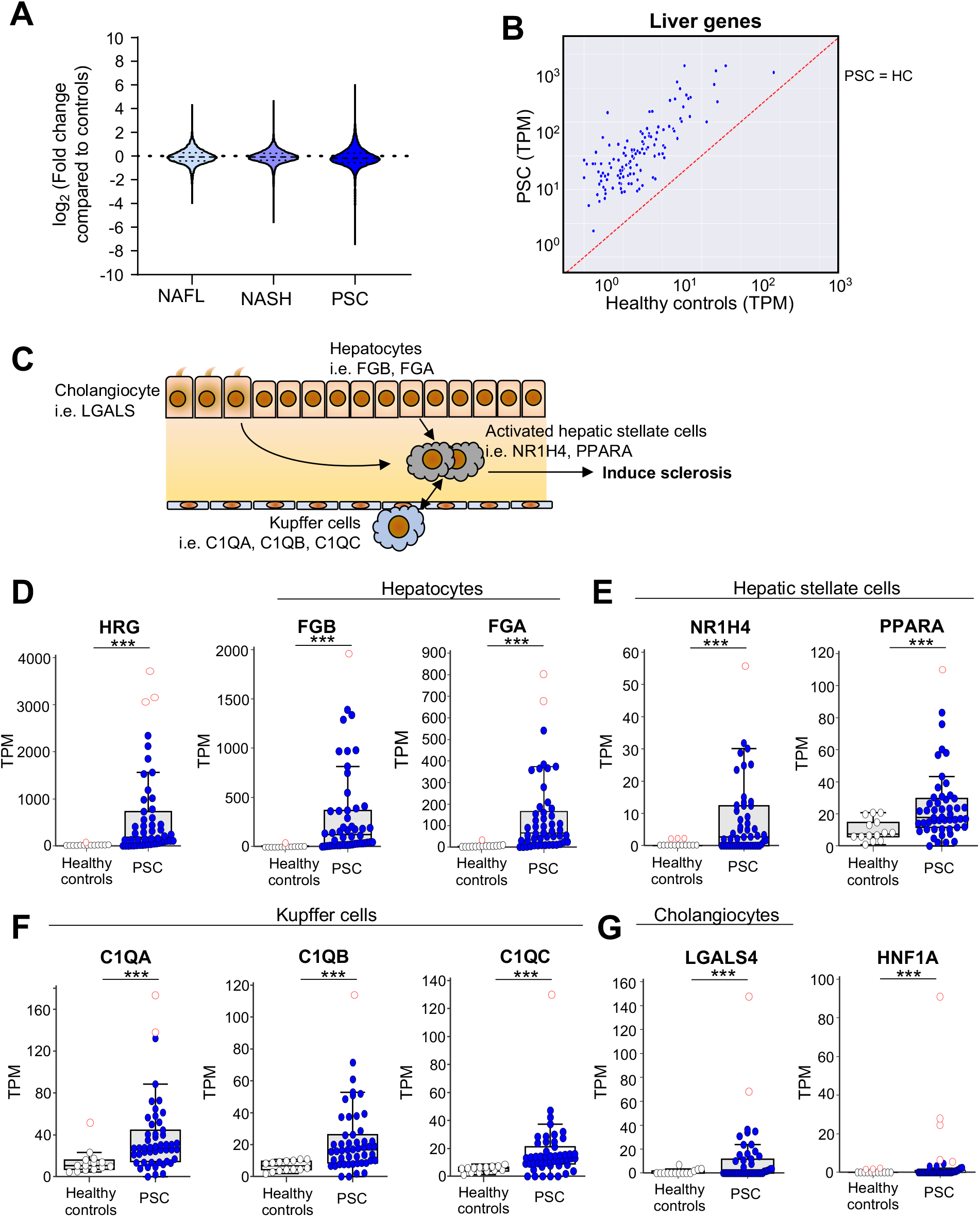
Cf-mRNA expression levels of liver cell type specific genes in subjects with PSC. (A) Fold change distribution of all genes in NAFL, NASH and PSC compared to healthy controls. (B) Gene expression correspondence of liver-specific genes between PSC and healthy controls (mean expression levels of individual genes were used for the correlation). (C) Schematics of liver cell type specific genes that may be involved in induction of sclerosis. (D) TPM levels of HRG, FGB and FGA genes between PSC and healthy controls. (*** *p* < 0.001) (E) TPM levels of hepatic stellate cells associated genes NR1H4 and PPARA genes between PSC and healthy controls. (*** *p* < 0.001) (F) TPM levels of Kupffer cells associated genes C1QA, C1QB and C1QC genes between PSC and healthy controls. (*** *p* < 0.001). (G) TPM levels of Cholangiocyte associated cells associated gene, LGALS4 and HNF1A between PSC and healthy controls. (*** *p* < 0.001).

### Characterization of PSC cf-mRNA transcriptome

To further interrogate the cf-mRNA transcriptomes of subjects with PSC, we used non-negative matrix factorization (NNMF) to examine whether the genes that are dysregulated in the circulation of this group are organized into functionally related clusters. We initially identified 12 gene clusters but eliminated clusters with small numbers of genes (less than 150 genes in the cluster) (Supplementary Fig. 8). The IPA pathway enrichment analysis of the remaining 7 individual clusters (Figure 4A) showed that several of the gene clusters were associated with biologically relevant processes of PSC, including the genes in Cluster 5 associated strongly with liver metabolism (Figure 4B) and the genes in Cluster 2 associated with integrin signaling *(38)*. Next, we used the GTEx database *(24)* to investigate whether these clusters have tissue- or cell type-specificity. Cluster 5 had high expression levels of liver-specific genes, but Cluster 1 had high expression levels of CD4 cells (Figure 4C, Supplementary Figure 10). A recent study identified that CD4 positive T cells in subjects with PSC exhibited reduced apoptosis and suppressed proapoptotic Bim in peripheral blood *(39)*. Consistently we observed that Cluster 1 was higher in expression in PSC compared to healthy control subjects (Figure 4D); we observed higher expression levels of liver-specific genes in PSC subjects in Cluster 5 and was higher in subjects with PSC compared to that of healthy controls (Figure 4D). Next, we evaluated whether these transcriptional clusters corresponded to specific PSC subject subsets. We performed unsupervised hierarchical clustering of 50 subjects with PSC based on the magnitude of the 7 clusters. Our analysis revealed that these gene clusters corresponded to five distinct subject groups with Clusters 1, 4 and 7 appearing to overlap to form a subject group (Figure 4E). We identified that group 5 (liver-genes) and group 1 (CD4 cells) had the most subjects. Although we do not have additional clinical data to evaluate whether these PSC subsets have associations with key clinical parameters of PSC, our study highlights the potential of cf-mRNA as a possible tool to characterize PSC patient endophenotypes noninvasively.

**Figure 4:**
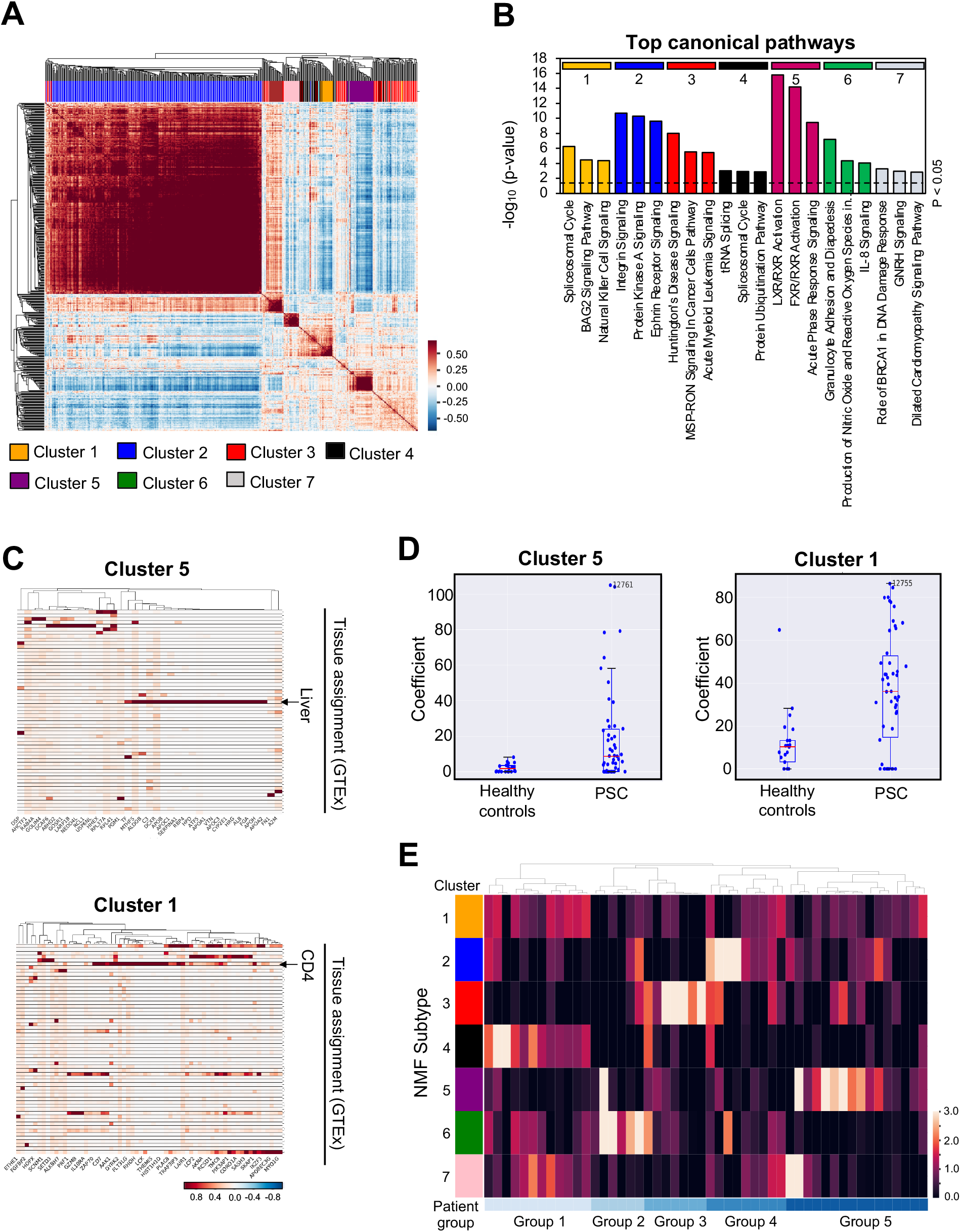
Identification PSC associated cf-mRNA gene-clusters and subject stratification using cf-mRNA gene clusters. (A) Consensus matrix NMF clustering of genes that are differentially expressed in cf-mRNA of subjects with PSC compared to healthy controls. 7 major cluster subtypes are labelled. (B) Most significant canonical pathway identified for each of 7 major clusters (IPA gene set enrichment analysis). (C) The expression of the Cluster 5 genes (total 161 genes) and Cluster 1 genes (total 356 genes) their tissue specific gene-expression using GTEx. (D) The individual levels of Cluster 5 and 1 coefficient between healthy controls and subjects with PSC. (E) Unsupervised clustering of subjects with PSC using their cf-mRNA profile based on NMF clusters identified in (A) revealed five groups of subjects with PSC.

In addition, we examined the key parameters of liver tests to ensure that the molecular characteristics of subjects with PSC are specific to PSC and not influenced by other liver abnormalities. As expected, mean levels for Aspartate transaminase (AST) and Alkaline phosphatase (ALP) of subjects with PSC were higher than the upper normal range of healthy individuals, while mean alanine aminotransferase (ALT) levels were within upper normal range for healthy individuals (Supplementary Figure 11A, B and C). Furthermore, since IBD is a frequent comorbidity of PSC, we examined whether the elevation of the circulating liver-specific cf-mRNA gene transcripts, Histidine rich glycoprotein (HRG) and Fibrinogen beta chain (FGB) as well as albumin (ALB), well-recognized highly expressed liver genes, were influenced by subjects’ IBD status. The levels of all three liver-specific genes did not differ between subjects with PSC with or without IBD, indicating that the elevation of liver-specific genes in these subjects is unlikely due to IBD (Supplementary Figure 11D).

### Diagnostic cf-mRNA classifiers for PSC

Considering that we have identified elevated levels of circulating liver cf-mRNA gene transcripts in subjects with PSC, we next investigated whether we can utilize these liver-specific genes to develop classifiers for PSC diagnosis. We used logistic regression analysis with repeated (15x) 2-fold cross-validation and tested genes that are specific to the liver. Liver-specific cf-mRNA transcripts alone differentiated subjects with PSC from healthy controls (AUC = 0.960: Figure 5A). However, when we generated a diagnostic classifier using all genes, the overall performance of the diagnostic classifier did not improve (AUC = 0.960: Figure 5B), indicating that the liver-genes contributed to the majority of the classifier performance. Since this classifier was derived only with the genes that are originated from the liver, the diagnostic classifier most likely captures the pathophysiological changes that are occurring in the liver.

**Figure 5:**
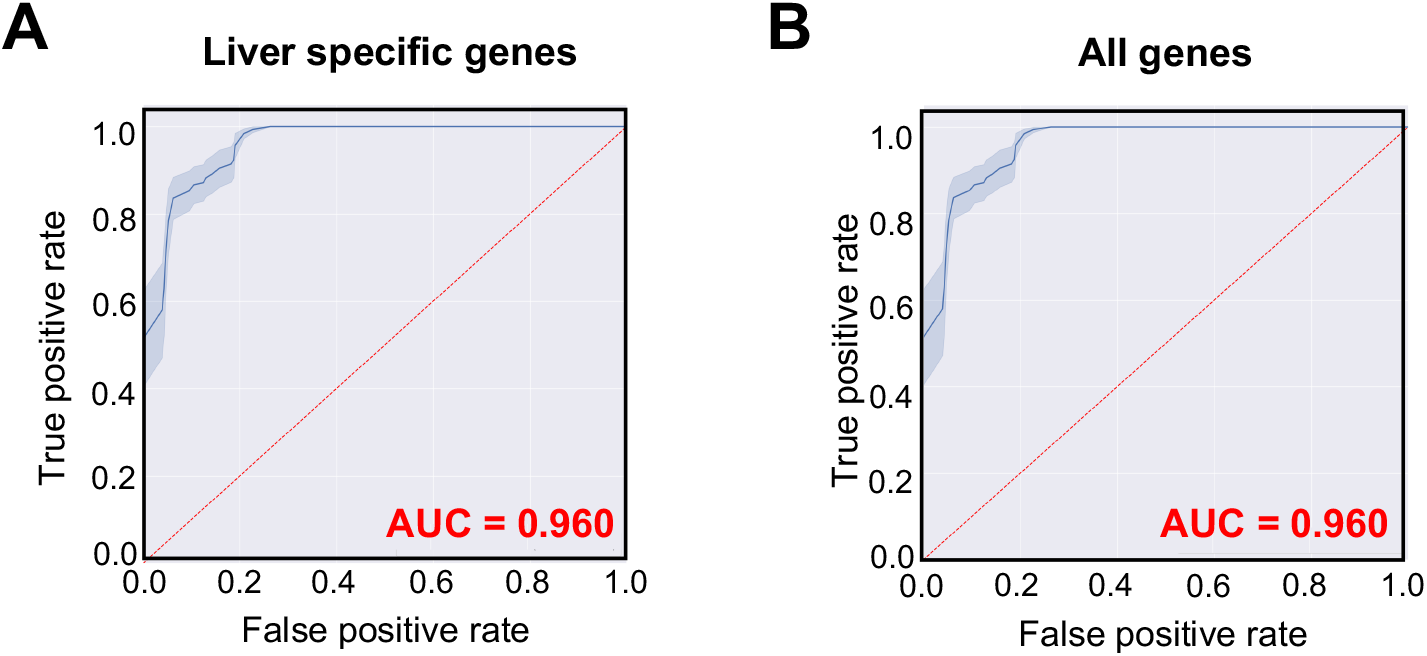
Establishment of PSC diagnosis classifiers. (A) ROC curve of cf-mRNA PSC classifiers using liver-specific genes as input comparing to healthy controls. (B) ROC curve of cf-mRNA PSC classifiers using all genes as input comparing to healthy controls.

Next, we evaluated whether building diagnostic classifiers between subjects with PSC and NAFL/NASH using only liver-specific cf-mRNA gene transcripts (Supplementary Fig. 12B) and all cf-mRNA gene transcripts (Supplementary Fig. 12B), to evaluate whether genes that are dysregulated in PSC differs from other liver diseases. The performance of the liver-specific cf-mRNA transcript classifiers to discriminate subjects with PSC from NAFL/NASH was reduced compared to that of healthy controls (NAFL: AUC = 0.831 and NASH: AUC= 0.801), most likely due to the similarities of underlying pathology of PSC and NAFL/NASH in comparison to healthy controls. In contrast, the addition of non-liver genes improved the overall performance of the diagnostic classifiers (NAFL: AUC = 0.862 and NASH: AUC= 0.849), indicating that the circulating non-liver gene transcripts may assist in distinguishing subjects with NAFL and NASH from PSC. Although these classifiers were derived from a relatively limited number of subject samples, our study highlights the potential clinical utility of cf-mRNA profiling in PSC diagnosis.

## DISCUSSION

PSC is a rare chronic disease of bile duct, with unclear etiology and pathogenesis. We conducted cf-mRNA profiling on serum samples of subjects with PSC and identified high levels of liver-specific cf-mRNA transcript dysregulation. We used multiple data sources to identify liver tissue- and cell type-specific cf-mRNA transcripts. These dysregulated cell free liver-specific gene transcripts appear to reflect functional changes expected to be associated with PSC pathogenesis. Utilization of NMF to dysregulated cf-mRNA in PSC resulted in identification of 7 major clusters and these clusters corresponded to 5 subject groups. We subsequently established cf-mRNA-based diagnostic classifiers for PSC using liver-specific transcripts.

Comparison of PSC and NAFL/NASH classifiers supported shared fractional underlying pathology and role of different non-liver transcripts for these two liver diseases.

One of the major challenges with developing nucleic acid-based blood biomarkers is the ambiguity of organ-specificity. The specificity of methylation of cfDNA has been proposed to indirectly infer the origin of the DNA *(40)*, though genomic methylation does not capture the epigenetic modifications of histones also known to be integrally involved in the specificity of gene expresion *(41)*. In contrast, RNA is a direct measure of gene expression and is a dynamic process in acute physiological events *(42, 43)*. Furthermore, multiple tissue sequencing datasets have identified a number of genes that are expressed specifically or are highly enriched in organs or cell types *(24-26, 33, 44-47)*. The assessment of cell free tissue-specific gene transcripts in the circulation allowed us to directly monitor changes occurring in the organ and cell type of interest. Cf-mRNA is harbored by extracellular lipid bilayer vesicles (eg. apoptotic bodies, exosomes and microvesicles), membrane-less complexes (e.g. extracellular particles, RNA granules, stress granules, and supermeres) and have a half-life similar to cfDNA. Studies have implicated these extracellular cf-mRNA carriers in intercellular communication to proximal and distal cells as well as potentially serving as biosensors for disruptions in cell homeostasis though additional definitive studies are needed *(48)*. In the present study, we showed that substantially higher levels of liver-specific gene transcripts were detected in the cf-mRNA of PSC compared to healthy controls as well as NAFLD/NASH. PSC is known to cause inflammation and scarring of the liver and subsequent accumulation of bile in the liver results in liver damage *(1)*. Interestingly, HRG, a liver-specific gene which is known to promote inflammation and fibrosis in the liver *(49)*, was the most highly elevated circulating transcripts in subjects with PSC and the circulating HRG protein has been shown to be elevated in subjects with mild liver cirrhosis *(50)*. Consistent with our findings, hepatotoxin-induced liver damage resulted in elevation of circulating cell free liver-specific mRNAs in mice *(51)*. Similarly, we observed upregulation of liver-specific genes that are known to be associated with PSC pathogenesis including, R1H4 and C1Q genes *(36, 37)*. Although we identified upregulation of liver-specific gene transcripts in another liver disease (NAFL and NASH), subjects with PSC had substantially higher levels of these gene transcripts. Interestingly, hepatocytes have been shown to excrete extracellular vesicles following liver injury to promote repair *(52)*, a possible mechanistic explanation for the elevation of liver-specific genes—in particular hepatocyte associated genes—in the circulation of subjects with PSC. While further investigation is required to understand the underlying causes of elevated liver-specific transcripts in the circulation of subjects with PSC, the present study highlights the potential utility of comprehensive molecular characterization of the bioactive cf-mRNA transcriptome for examining the transcriptional changes in the liver.

Unlike other liver diseases, in the presence of abnormal cholangiogram, liver biopsy is not performed for diagnosis for large duct PSC. A previous retrospective study with 138 choloangiographically confirmed subjects with PSC indicated that liver biopsy seldom adds useful information for PSC diagnosis *(53)*. Moreover, in a previous attempt to develop a PSC risk stratification model, the inclusion of biopsy was abandoned due to its invasive nature and inherent sample variability *(54, 55)*. In the present study, we utilized a cf-mRNA sequencing and machine learning platform to conduct comprehensive transcriptomic profiling of subjects with PSC to capture the entire cf-mRNA secretome. We identified distinct molecular characteristics in the cf-mRNA transcriptome including evaluation of key genes that are associated with specific liver cell types. Recapitulation of expected cell types increases the confidence in the study results. The non-invasive nature of blood-based molecular profiling provides significant potential for diseases such as PSC, where therapeutic development is lagging, and clinical sampling is challenging. Beyond use in pre-clinical studies focused on evaluation of pharmacotherapy target engagement and off target disruptions, comprehensive non-invasive molecular profiling could be utilized for patient pre-selection for drugs or combinations that target specific pathways and used to monitor therapeutic efficacy and safety. Moreover, we demonstrated that not only can we discriminate subjects with PSC from healthy controls. Further investigation, validation, and optimization using larger sample sets will be important to translate these promising findings into clinical care.

Limitations of the study include the small number of subjects, the use of only cross validation rather than a fully independent test set and the sourcing of healthy controls and liver disease samples from different institutions. An additional limitation is the use of opportunistic archival sera samples. In addition, the highly heterogeneous extracellular vesicles and particles precludes purification of any specific type of complex preventing identification of the specific vesicles and particles that are the origin of cf-mRNA *(48, 56)*. The extracellular vesicle database used likely includes RNA from a diverse set of extracellular complexes so attribution to a specific extracellular complex or biogenesis route is not possible. Our study captures the entire cf-mRNA secretome and therefore may be more inclusive than studies that attempt enrichment or purification.

In conclusion, we conducted comprehensive noninvasive cf-mRNA profiling in serum samples obtained from subjects with PSC and identified elevated levels of circulating liver-specific transcripts consistent with present understanding of PSC pathogenesis. Continued review of liver specific and non-liver specific transcripts in cf-mRNA studies may uncover additional underlying pathologies beyond those understood for PSC. We further developed preliminary cf-mRNA classifiers for the potential diagnosis of PSC. Our results highlighted the potential of cf-mRNA profiling as a platform for non-invasive molecular characterization of liver diseases such as PSC and a tool for the development of non-invasive disease biomarkers.

## MATERIALS AND METHODS

### Clinical specimens

We examined total of 50 PSC serum specimens from Indiana University, IN and 20 healthy control serum specimens from San Diego Blood Bank, CA. The diagnosis of PSC was established with MRCP/ERCP. We used additional 235 biopsy-proven NAFL (steatosis without hepatocellular ballooning) and NASH (steatosis with hepatocellular ballooning, lobular inflammation with or without fibrosis) samples collected from Indiana University. The sequencing data of NAFL and NASH have been published previously and are available on following database (SRA: PRJNA701722) *(18)*. The detailed subject demographics and clinicopathological characteristics are shown in Supplementary Table 1. Written informed consent was obtained from all subjects, and the study was approved by the institutional review boards of all the participating institutions.

### RNA extraction, library preparation and whole-transcriptome RNA-seq

RNA was extracted from up to 1 mL of serum using QIA amp Circulating Nucleic Acid Kit (Qiagen) and eluted in 15 µl volume. ERCC RNA Spike-In Mix (Thermo Fisher Scientific, Cat. # 4456740) was added to RNA as an exogenous spike-in control according to manufacturer’s instruction (Ambion). Agilent RNA 6000 Pico chip (Agilent Technologies, Cat. # 5067-1513) was used to assess the integrity of extracted RNA. RNA samples were converted into a sequencing library as described previously *(17)*. Qualitative and quantitative analysis of the NGS library preparation process was conducted using a chip-based electrophoresis and libraries were quantified using a qPCR-based quantification kit (Roche, Cat. # KK4824). Sequencing was performed using Illumina NextSeq500 platform (Illumina Inc), using paired-end sequencing, 75-cycle sequencing. Base-calling was performed on an Illumina BaseSpace platform (Illumina Inc), using the FASTQ Generation Application. For sequencing data analysis, adaptor sequences were removed, and low-quality bases were trimmed using cutadapt (v1.11). Reads shorter than 15 base-pairs were excluded from subsequent analysis. Read sequences greater than 15 base-pairs were compared to the human reference genome GRCh38 using STAR (v2.5.2b) with GENCODE v24 gene models. Duplicated reads were removed using the samtools (v1.3.1) rmdup command. Gene-expression levels were calculated from de-duplicated BAM files using RSEM (v1.3.0).

### Liver tissue and cell type specific gene establishment

GTEx Liver-specific genes are defined as genes that show substantially higher expression in a particular tissue compared to other tissue types. Tissue-specific transcriptome expression levels were obtained from the following two public databases: GTEx (https://www.gtexportal.org/home/) for gene expression across 51 human tissues *(24)* and Blueprint Epigenome (http://www.blueprint-epigenome.eu/) for gene expression across 56 human hematopoietic cell types. For each individual gene, the tissues were ranked by their expression of that particular gene and if the expression in the top tissue (cell-type) was > 20-fold higher than all the other tissues, the gene was considered specific to the top tissue. The list of liver genes is shown in Supplementary Table 2. Furthermore, “elevated genes” in the liver tissue from Human Protein Atlas database (downloaded on March 15^th^, 2022) *(25)* were used as the HPA liver-specific genes. For cell type specific genes, the PanglaoDB database was used to categorize liver cell type specific genes *(26)*.

### Bioinformatic analysis/Classifier development

We used logistic regression analysis with L1 regularization within the scikit-learn Python library for implementation of the classification. Meta-parameters were determined by repeated (15x) cross-validation, by randomly withholding 40% of the samples for validation within the “training cohort”.

### Computational Deconvolution analysis using non-negative matrix factorization (NMF)

A normalization was first implemented whereby the expression levels of each gene were divided by its maximum value across the samples. This step is designed to rescale the expression levels among different genes to avoid a few highly expressed genes dominating the decomposition process. The normalized expression matrix was then subject to NMF decomposition using sklearn.decomposition.NMF within the Python library Scikit-learn (https://scikit-learn.org/stable/). NMF decomposition achieves a more parsimonious representation of the data by decomposing expression matrix into the product of two matrices X = WH. X is the expression matrix with n rows (n samples) and m columns (m genes); W is the coefficient matrix with n rows (n samples) and p columns (p components); H is the loading matrix with p rows (p components) and m columns (m genes). W is in a sense a summarization of the original matrix H with reduced number of dimensions. H contains information about how much each gene contribute to the components. Biological interpretation of the derived components was achieved by performing pathway analysis on the top genes that contribute the most to each component. We performed subject grouping by performing hierarchical clustering on the coefficient matrix W. Hierarchical clustering was implemented using Python library SciPy (v1.3.0) class scipy.cluster.hierarchy.linkage with parameters method = “average” and metric = “correlation”. A small offset (10^−5^) was added to all values to avoid issues with zeroes.

### Statistical analysis

Demographic variables were summarized using mean and standard deviation or frequency and percentage. Risk scores derived from the gene-classifier multivariate logistic regression model were used to plot receiver-operating-characteristic (ROC) curves and calculate area under the curves (AUCs). The limited number of subjects with PSC prevented creation of a separate validation data set. Therefore, repeated (15x) 2-fold cross-validation was used to assess potential out of sample performance variation of the AUC. Assessments involving two groups were evaluated using Student’s t-test, while assessments involving more than two groups were evaluated using analysis of variance (ANOVA). All statistical analyses were performed using Python version 2.7.5 *(57)*, R version 3.6.0 *(58)*. URL https://www.R-project.org/. and MedCalc version 19 (Medcalc Software bvba, Ostend, Belgium).

## Data Availability

All data produced in the present study are available upon reasonable request to the authors

## ACKNOWLEDGEMENT

We thank Kayla Gelow and Emily R Smith for sample collection and data management.

## AUTHOR CONTRIBUTIONS

N.C., M.N. and S.T., conceived and designed research; J.Z., and J.V.B and S.T., analyzed data; N.C., R.V., C.L., S.G., J.V.B., J.Z., A.I, D.A.R., M.N., S.R.Q., J.J.S. and S.T. interpreted results of experiments; J.Z., J.V.B. and S.T. prepared figures; N.C., J.J.S. and S.T. drafted manuscript; N.C., R.V., C.L., S.G., J.V.B., J.Z., A.I, D.A.R., M.N., S.R.Q., J.J.S. and S.T. edited and revised manuscript.

