## Supplementary Material for "Circulating cell-free messenger RNA secretome characterization of primary sclerosing cholangitis"

**Supplementary Figure 1**

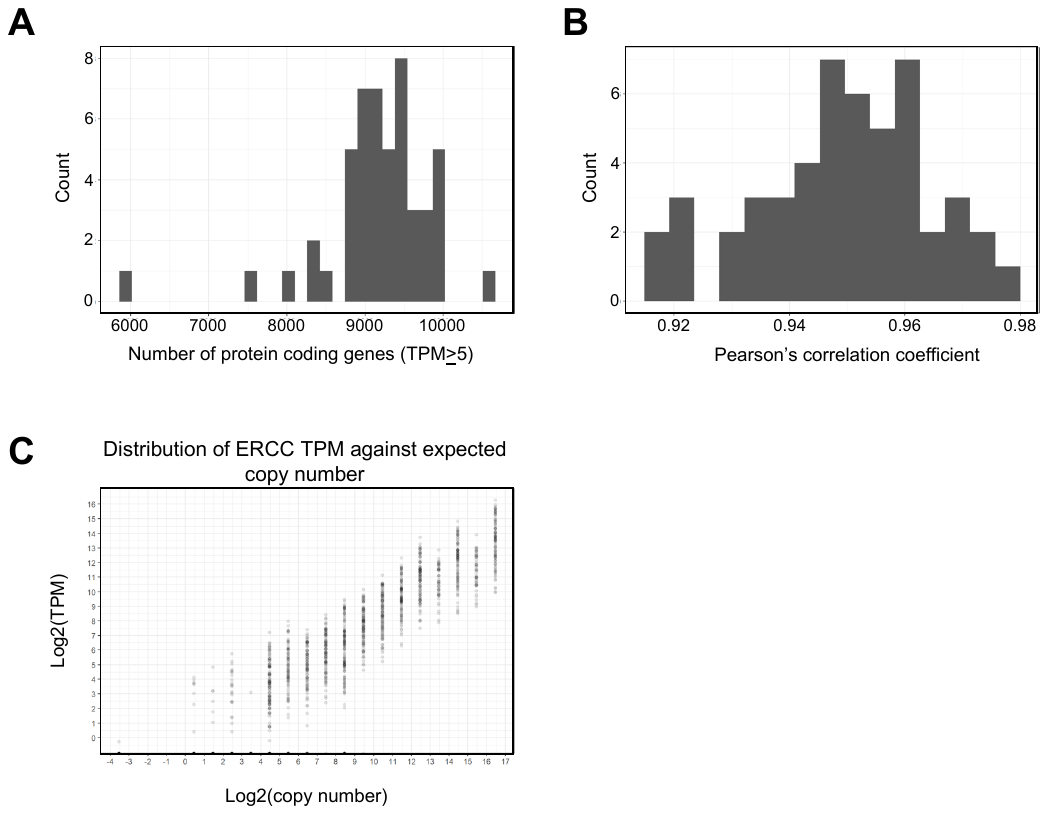

**Supplementary Figure 1:** Cf-mRNA Seq is a robust approach for the characterization of cf-mRNA transcriptome. (A) Histogram of genes detected per sample (TPM > 5). (B) Histogram of Spearman’s correlation coefficient of observed versus expected copy number based on spiked-in ERCC control.

**Supplementary Figure 2**

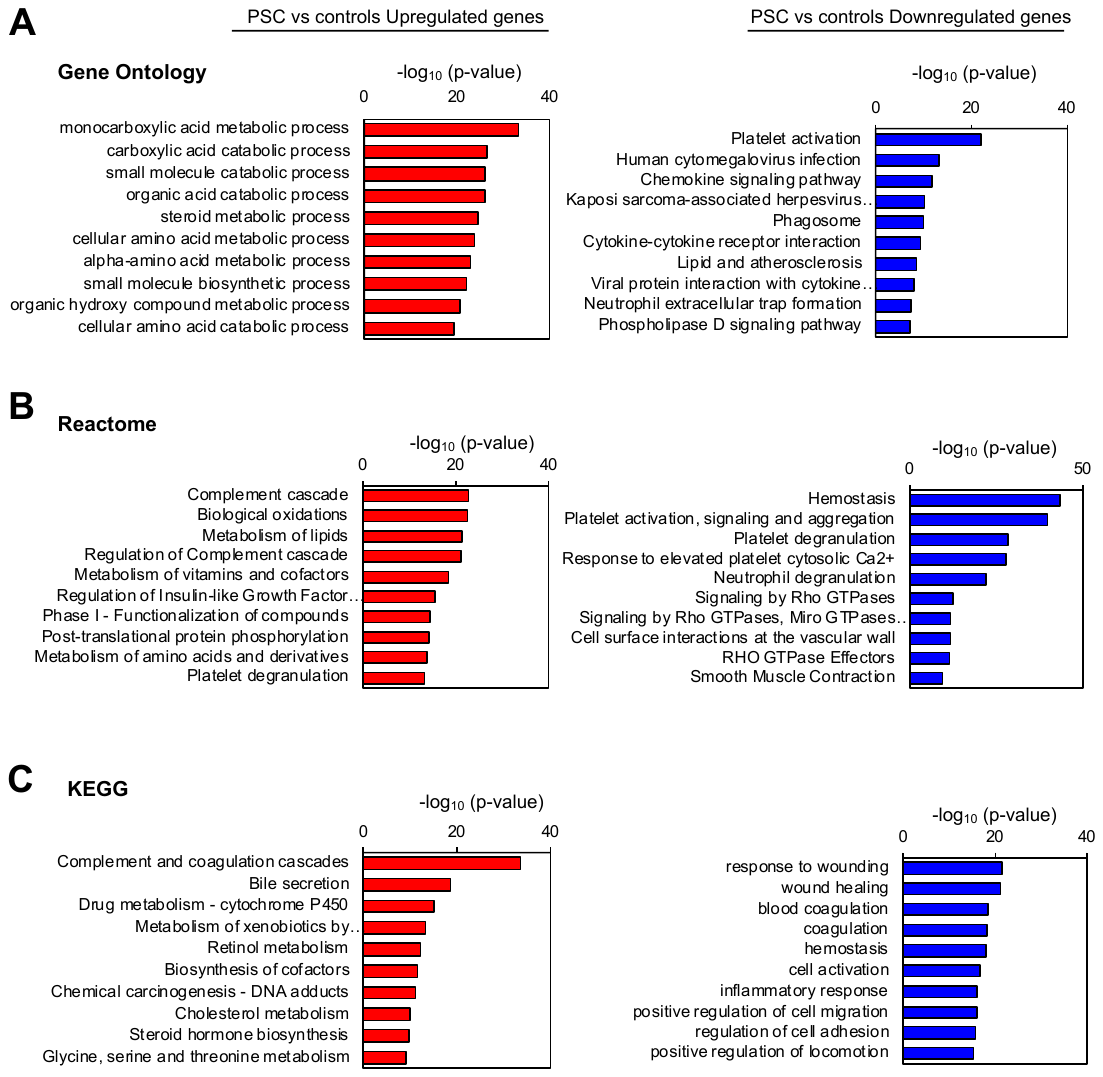

**Supplementary Figure 2:** Pathways associated with PSC associated cf-mRNA transcripts. (A) Most significantly enriched pathways determined by Gene Ontology using differentially expressed genes (PSC vs controls) as input (Top). Upregulated pathways (Left) and downregulated pathways (Right). (B) Reactome pathway analysis using differentially expressed genes (PSC vs controls) as input (Middle). Upregulated pathways (Left) and downregulated pathways (Right). (C) KEGG pathway analysis using differentially expressed genes (PSC vs controls) as input (Bottom). Upregulated pathways (Left) and downregulated pathways (Right).

**Supplementary Figure 3**

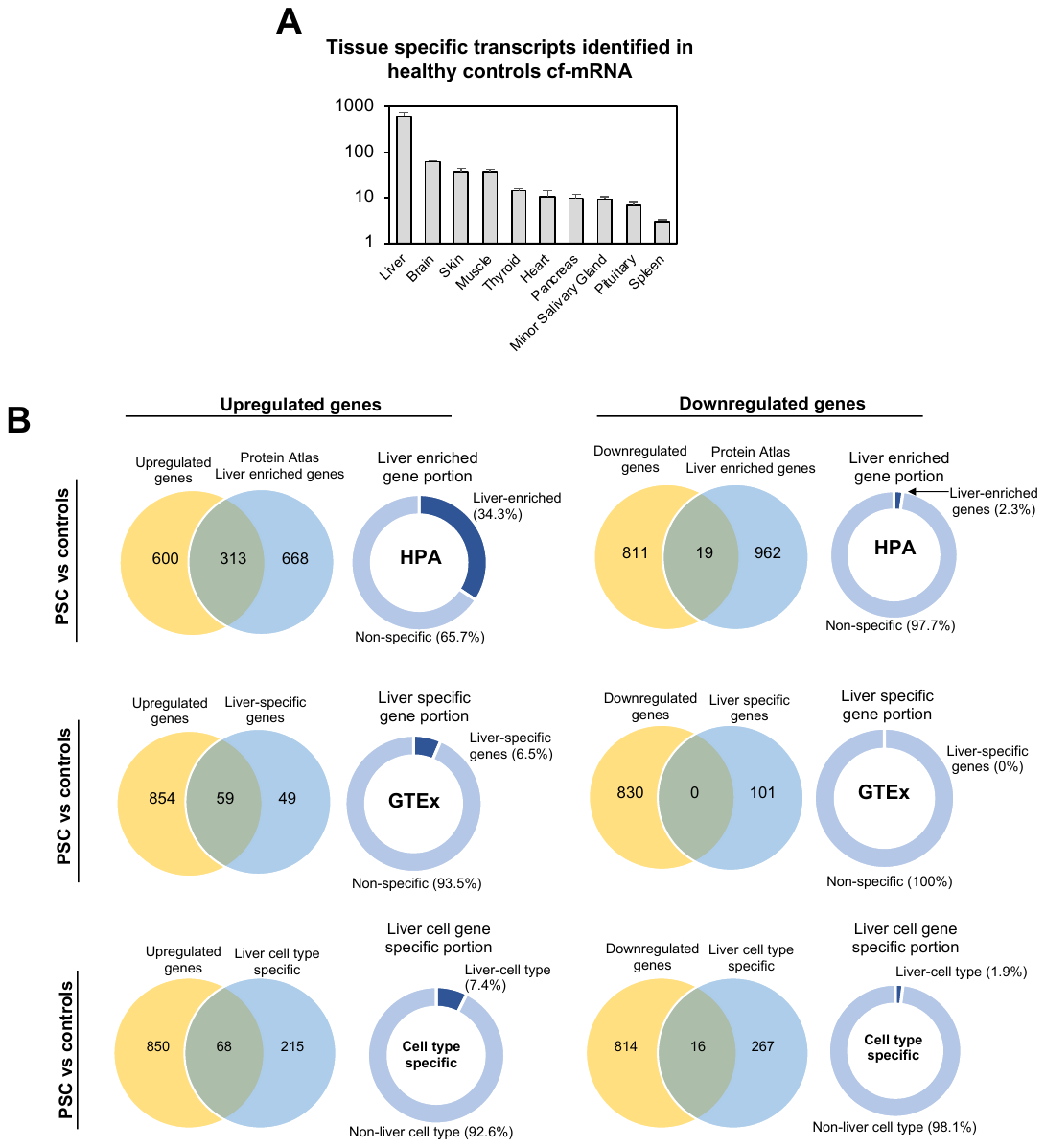

**Supplementary Figure 3:** Summary of tissue specific genes for healthy controls and quantification of liver tissue specific and cell type specific genes that are dysregulated in PSC. (A) Quantification of tissue specific transcripts identified in cf-mRNA of healthy controls (Mean ± SEM). (B) Quantification of liver tissue and cell type specific genes in genes that are differentially expressed in subjects with PSC compared to healthy controls (upregulated genes, left; downregulated genes, right). Reference data sets used to calculate the liver tissue and cell type specificities are: Human Protein Atlas (HPA) (top), GTEx (middle) and PanglaoDB (bottom). Overlap between upregulated genes and each liver tissue or cell type specific references. Percentage of liver specific genes in upregulated genes. Overlap between downregulated genes and each liver tissue or cell type specific references. Percentage of liver specific genes in downregulated genes (left to right).

**Supplementary Figure 4**

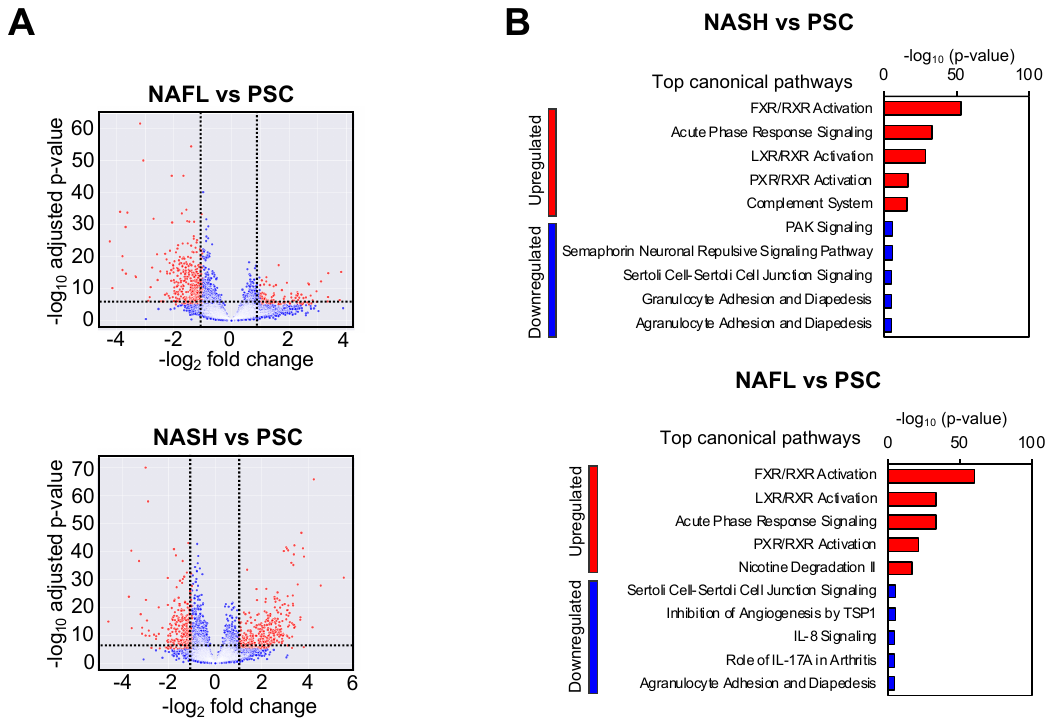

**Supplementary Figure 4:** Genes and pathways that are dysregulated in subjects with PSC compared to NAFL/NASH. (A) Volcano plots showing dysregulation of cf-mRNAs in subjects with PSC compared to that of NAFL (top) and NASH (bottom). (B) (C) Top IPA canonical pathways for genes that are dysregulated in PSC, upregulated (top) and downregulated (bottom) genes are used as input.

**Supplementary Figure 5**

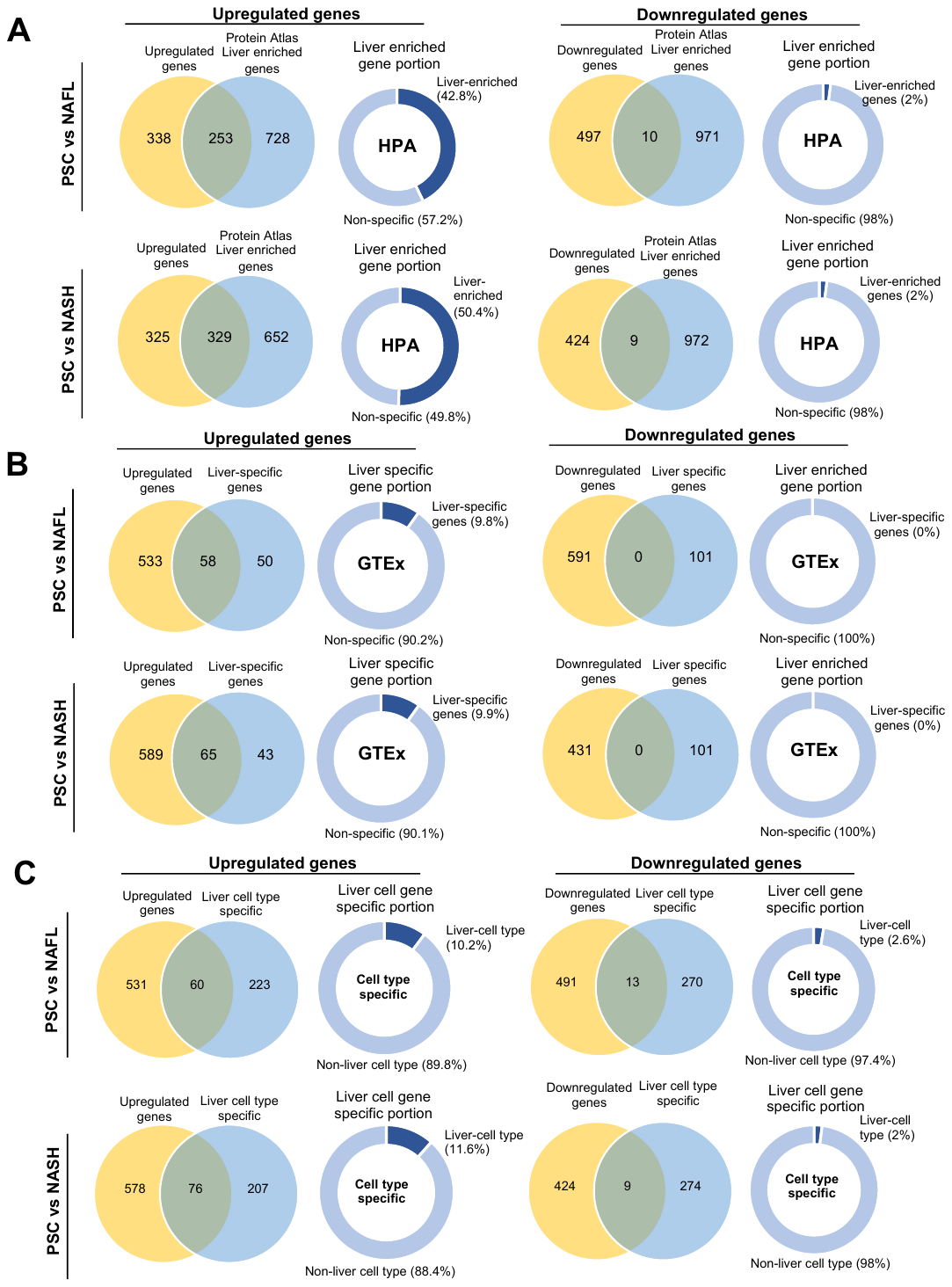

**Supplementary Figure 5:** Quantification of liver tissue specific and cell type specific genes that are dysregulated in PSC compared to NAFL and NASH. (A) Quantification of liver specific genes in genes that are differentially expressed in subjects with PSC compared to NAFL (top) and NASH (bottom) using Human Protein Atlas (HPA) as the reference (upregulated genes, left; downregulated genes, right). (B) Quantification of liver specific genes in genes that are differentially expressed in subjects with PSC compared to NAFL (top) and NASH (bottom) using GTEx as the reference (upregulated genes, left; downregulated genes, right). (C) Quantification of liver cell type specific genes in genes that are differentially expressed in subjects with PSC compared to NAFL (top) and NASH (bottom) using PanglaoDB as the reference (upregulated genes, left; downregulated genes, right). Overlap between upregulated genes and each liver tissue or cell type specific references. Percentage of liver specific genes in upregulated genes. Overlap between downregulated genes and each liver tissue or cell type specific references. Percentage of liver specific genes in downregulated genes (left to right).

**Supplementary Figure 6**

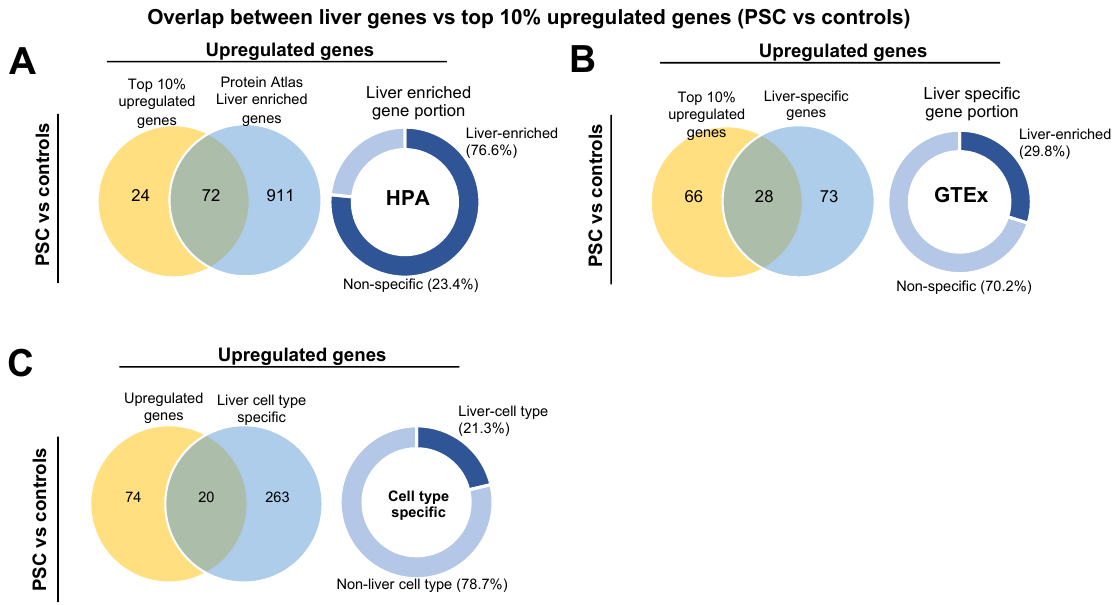

**Supplementary Figure 6:** Quantification of liver tissue specific and cell type specific genes in top 10% of genes that are upregulated in PSC compared to controls. (A) Quantification of liver specific genes in genes that are upregulated (top 10%) in subjects with PSC compared to controls using Human Protein Atlas (HPA) as the reference (Overlap between 10% upregulated and HPA liver specific genes, left; Percentage of liver specific genes in top 10% upregulated genes). (B) Quantification of liver specific genes in genes that are upregulated (top 10%) in subjects with PSC compared to controls using GTEx as the reference (Overlap between 10% upregulated and GTEx liver specific genes, left; Percentage of liver specific genes in top 10% upregulated genes). (C) Quantification of liver cell type specific genes in genes that are upregulated (top 10%) in subjects with PSC compared to controls using PanglaoDB as the reference (Overlap between 10% upregulated and PanglaoDB liver cell type specific genes, left; Percentage of liver cell type specific genes in top 10% upregulated genes).

**Supplementary Figure 7**

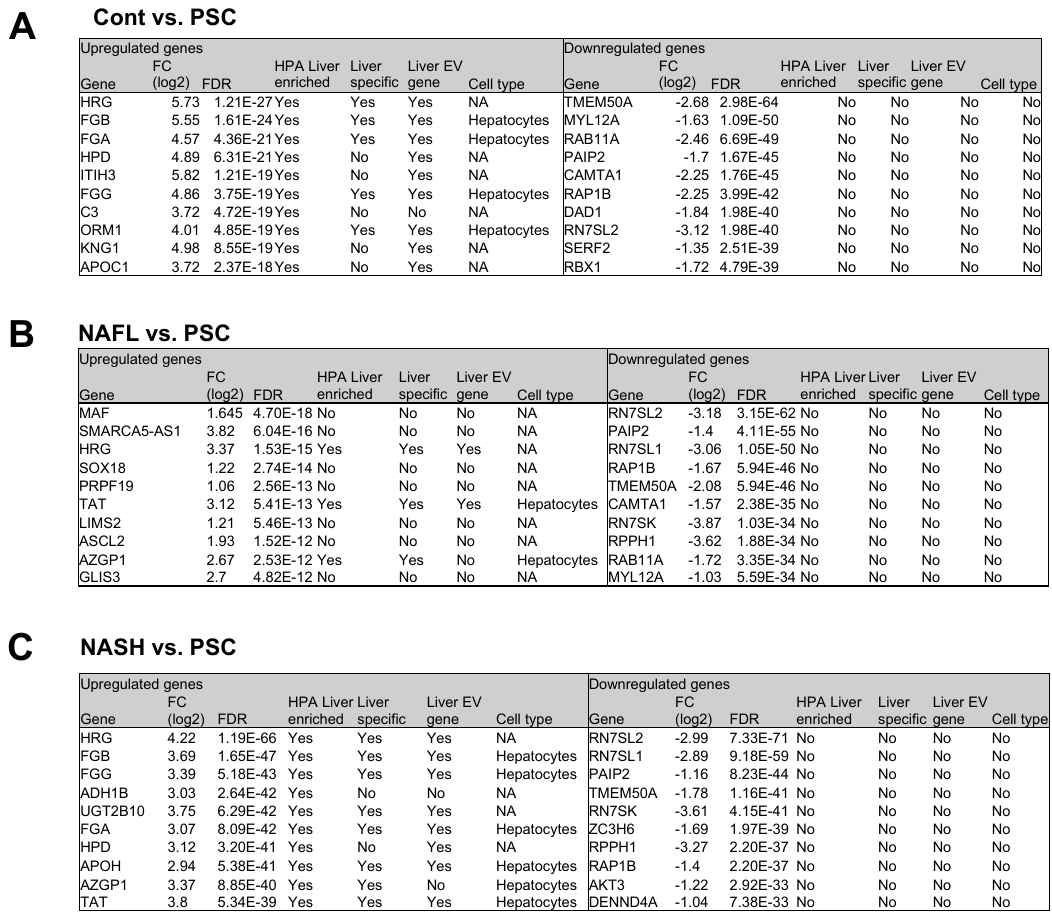

**Supplementary Figure 7:** Evaluation of liver specificity in top 10 dysregulated genes in PSC compared to healthy controls, NAFL and NASH (A) A table summarizing the liver associated molecular characteristics of the top 10 most upregulated genes in cf-mRNA of subjects with PSC compared to healthy controls (upregulated, left; downregulated, right). (B) A table summarizing the liver associated molecular characteristics of the top 10 most upregulated genes in cf-mRNA of subjects with PSC compared to NAFL (upregulated, left; downregulated, right). (C) A table summarizing the liver associated molecular characteristics of the top 10 most upregulated genes in cf-mRNA of subjects with PSC compared to NASH (upregulated, left; downregulated, right).

**Supplementary Figure 8**

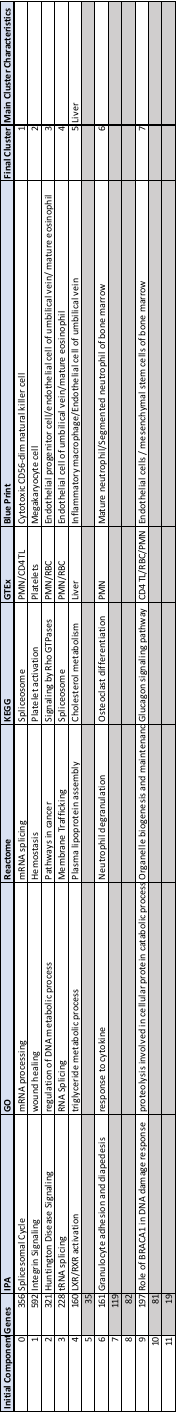

**Supplementary Figure 8:** Table summarizing the characteristics of each NMF clusters identified in Figure 3A and B.

**Supplementary Figure 9**

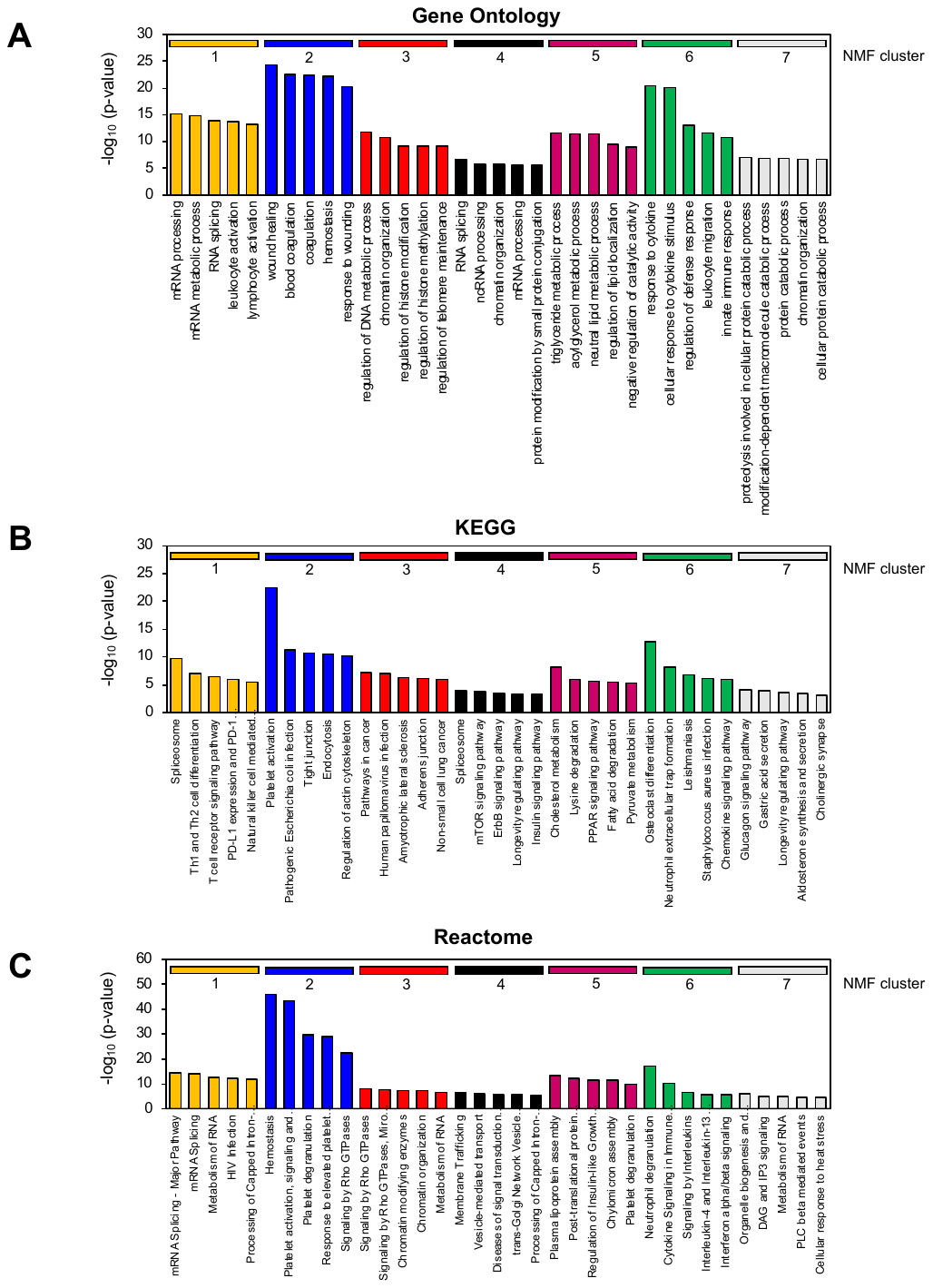

**Supplementary Figure 9:** Prominent signaling pathways of NMF clusters. (A) Most significant pathway identified for each of 7 major clusters (Gene Ontology enrichment analysis). (B) Most significant pathway identified for each of 7 major clusters (Reactome enrichment analysis). (C) Most significant pathway identified for each of 7 major clusters (KEGG enrichment analysis).

**Supplementary Figure 10**

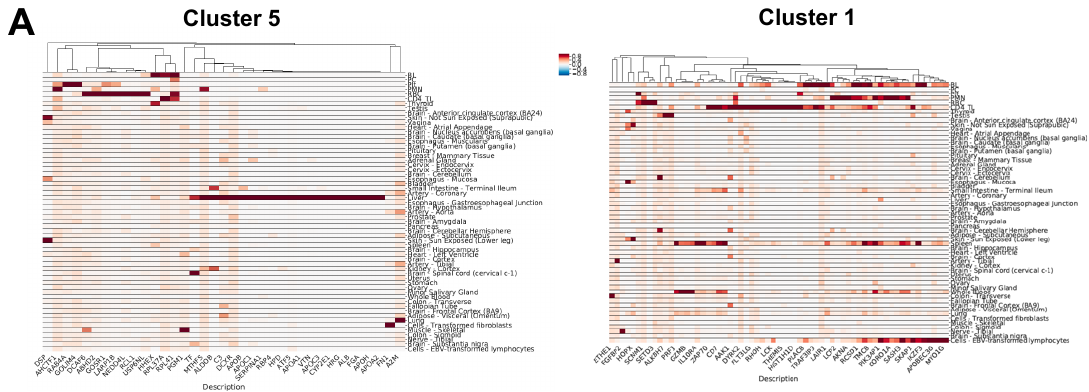

**Supplementary Figure 10:** Evaluation of tissue specificities in genes in each NMF cluster. (A) Tissue specificity of Cluster 5 genes using GTEx as the reference. (B) Tissue specificity of Cluster 1 genes using GTEx as the reference.

**Supplementary Figure 11**

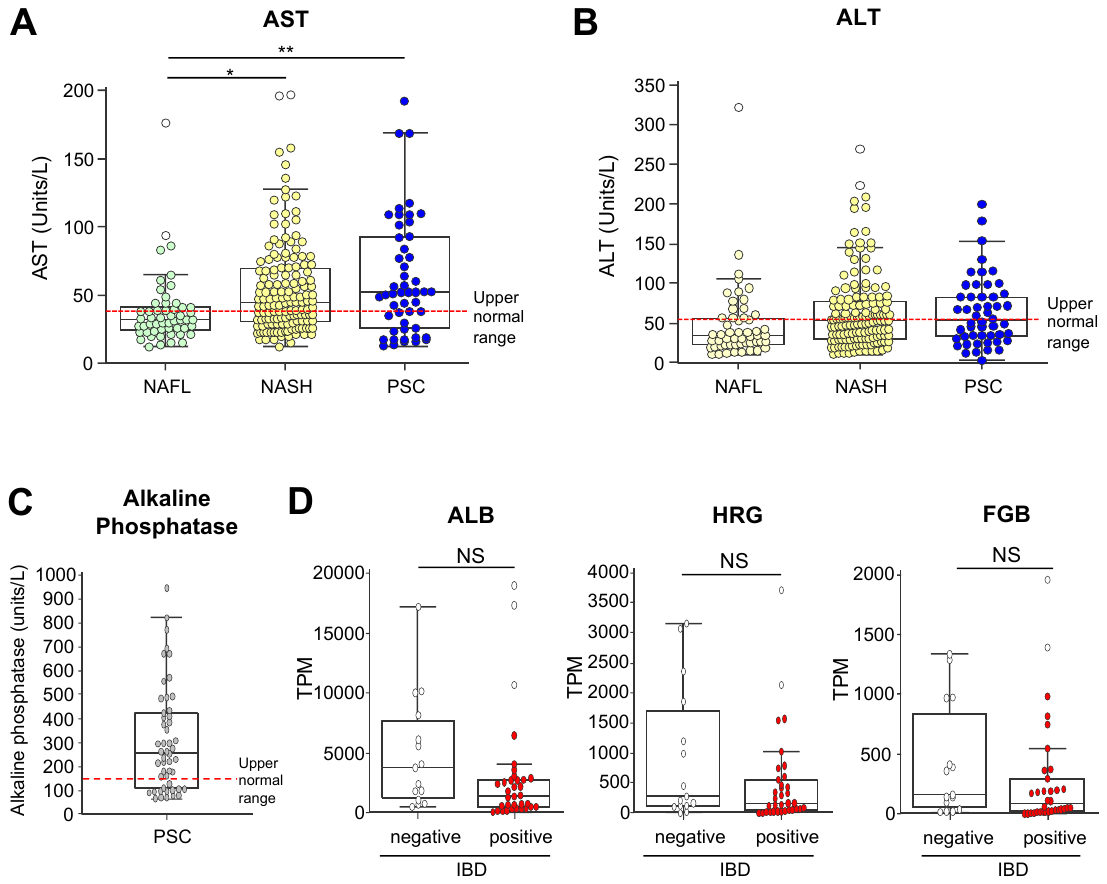

**Supplementary Figure 11:** AST and ALT levels and association of circulating liver-specific genes to IBD status. (A) AST levels in subjects with liver diseases. Dotted line represents upper limit for the normal range. (B) ALT levels in subjects with liver diseases. Dotted line represents upper limit for the normal range. (C) Alkaline phosphatase levels in subjects with PSC. Dotted line represents upper limit for the normal range. (D) The levels of liver-specific transcripts in subjects with or without IBD. ALB (left), HRG (center) and FGB (right).

**Supplementary Figure 12**

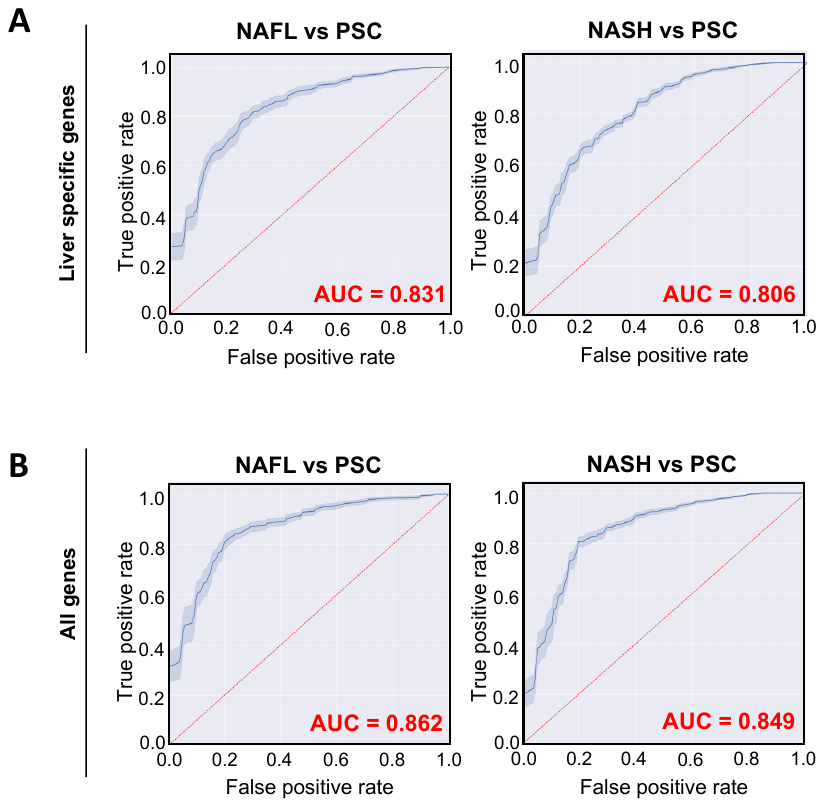

**Supplementary Figure 12:** Evaluation of PSC diagnosis classifiers efficacy for PSC vs NAFL or NASH. (A) ROC curve of cf-mRNA PSC classifiers using liver-specific genes as input comparing to healthy controls (PSC vs NAFL, left; PSC vs NASH, right) (B) ROC curve of cf-mRNA PSC classifiers using all genes as input comparing to healthy controls (PSC vs NAFL, left; PSC vs NASH, right).

**Supplementary Table 1:** Subject characteristic table

| **Disease** | **PSC** | **NAFL** | **NASH** | **Healthy controls** |
| --- | --- | --- | --- | --- |
| n | 50 | 52 | 164 | 20 |
| Sex (m) | NA | 22 | 49 | 12 |
| Age | NA | 48.2 ± 1.8 | 50.5 ± 0.9 | 50.1 ± 4.1 |
| AST | 62.0 ± 6.1 | 38.5 ± 4.0 | 55.2 ± 2.7 | - |
| ALT | 64.0 ± 6.1 | 47.9 ± 7.3 | 62.9 ± 3.8 | - |
| Alk Phos | 308.5 ± 31.1 |  |  | - |
| Tbili | 1.9 ± 0.4 |  |  | - |
| IBD (Yes) | 35 (70%) |  |  | - |

**Supplementary Table 2:** List for liver-specific genes

| ABCG5 | C8G | GC | RP11-622A1.2 |
| --- | --- | --- | --- |
| AC003988.1 | C9 | GLYATL3 | RP11-685F15.1 |
| AC006037.2 | CFHR1 | HAO1 | RP11-753B14.1 |
| AC068535.3 | CFHR2 | HP | SAA4 |
| ACOT12 | CFHR3 | HPX | SERPINA10 |
| ADH1A | CFHR4 | HRG | SERPINA11 |
| ADH4 | CFHR5 | HULC | SERPINA6 |
| AFM | COLEC10 | IGFBP1 | SERPINA7 |
| AGXT | CPB2 | INHBC | SERPINC1 |
| AHSG | CRP | INHBE | SERPIND1 |
| AKR1C4 | CTD-2526M8.2 | ITIH1 | SLC10A1 |
| ALB | CTD-2582M21.1 | ITIH2 | SLC13A5 |
| AMBP | CTD-3162L10.4 | ITIH4 | SLC17A2 |
| ANGPTL3 | CYP2A6 | LBP | SLC25A47 |
| APCS | CYP2C8 | LPA | SLCO1B1 |
| APOA2 | CYP2E1 | MAT1A | SLCO1B3 |
| APOA5 | CYP8B1 | MBL2 | TAT |
| APOC2 | F13B | OR10J5 | TDO2 |
| APOC4 | F2 | ORM1 | U91324.1 |
| APOF | F7 | ORM2 | UGT1A4 |
| APOH | F9 | PLG | UGT2B10 |
| ASGR1 | FAM99A | PON1 | UGT2B4 |
| ASGR2 | FGA | PROC | UROC1 |
| BAAT | FGB | RDH16 |  |
| C8A | FGF21 | RP11-101E14.3 | |
| C8B | FGG | RP11-1151B14.2 | |
